# Performance Drift in a Mortality Prediction Algorithm during the SARS-CoV-2 Pandemic

**DOI:** 10.1101/2022.02.28.22270996

**Authors:** Ravi B. Parikh, Yichen Zhang, Corey Chivers, Katherine R. Courtright, Jingsan Zhu, Caleb M. Hearn, Amol S. Navathe, Jinbo Chen

## Abstract

**Research Objective:** Health systems use clinical predictive algorithms to allocate resources to high-risk patients. Such algorithms are trained using historical data and are later implemented in clinical settings. During this implementation period, predictive algorithms are prone to performance changes (“drift”) due to exogenous shocks in utilization or shifts in patient characteristics. Our objective was to examine the impact of sudden utilization shifts during the SARS-CoV-2 pandemic on the performance of an electronic health record (EHR)-based prognostic algorithm.

**Study Design:** We studied changes in the performance of Conversation Connect, a validated machine learning algorithm that predicts 180-day mortality among outpatients with cancer receiving care at medical oncology practices within a large academic cancer center. Conversation Connect generates mortality risk predictions before each encounter using data from 159 EHR variables collected in the six months before the encounter. Since January 2019, Conversation Connect has been used as part of a behavioral intervention to prompt clinicians to consider early advance care planning conversations among patients with ≥10% mortality risk. First, we descriptively compared encounter-level characteristics in the following periods: January 2019–February 2020 (“pre-pandemic”), March-May 2020 (“early-pandemic”), and June–December 2020 (“later-pandemic”). Second, we quantified changes in high-risk patient encounters using interrupted time series analyses that controlled for pre-pandemic trends and demographic, clinical, and practice covariates. Our primary metric of performance drift was false negative rate (FNR). Third, we assessed contributors to performance drift by comparing distributions of key EHR inputs across periods and predicting later pandemic utilization using pre-pandemic inputs.

**Population Studied:** 237,336 in-person and telemedicine medical oncology encounters.

**Principal Findings:** Age, race, average patient encounters per month, insurance type, comorbidity counts, laboratory values, and overall mortality were similar among encounters in the pre-, early-, and later-pandemic periods. Relative to the pre-pandemic period, the later-pandemic period was characterized by a 6.5-percentage-point decrease (28.2% vs. 34.7%) in high-risk encounters (p<0.001). FNR increased from 41.0% (95% CI 38.0-44.1%) in the pre-pandemic period to 57.5% (95% CI 51.9-63.0%) in the later pandemic period. Compared to the pre-pandemic period, the early and later pandemic periods had higher proportions of telemedicine encounters (0.01% pre-pandemic vs. 20.0% early-pandemic vs. 26.4% later-pandemic) and encounters with no preceding laboratory draws (17.7% pre-pandemic vs. 19.8% early-pandemic vs. 24.1% later-pandemic). In the later pandemic period, observed laboratory utilization was lower than predicted (76.0% vs 81.2%, p<0.001). In the later-pandemic period, mean 180-day mortality risk scores were lower for telemedicine encounters vs. in-person encounters (10.3% vs 11.2%, p<0.001) and encounters with no vs. any preceding laboratory draws (1.5% vs. 14.0%, p<0.001).

**Conclusions:** During the SARS-CoV-2 pandemic period, the performance of a machine learning prognostic algorithm used to prompt advance care planning declined substantially. Increases in telemedicine and declines in laboratory utilization contributed to lower performance.

**Implications for Policy or Practice:** This is the first study to show algorithm performance drift due to SARS-CoV-2 pandemic-related shifts in telemedicine and laboratory utilization. These mechanisms of performance drift could apply to other EHR clinical predictive algorithms. Pandemic-related decreases in care utilization may negatively impact the performance of clinical predictive algorithms and warrant assessment and possible retraining of such algorithms.

## INTRODUCTION

Clinical predictive models that rely on electronic health record (EHR) inputs, such as encounters, administrative codes, and laboratory values,^1^ are increasingly used in health care settings to direct resources to high-risk patients. Such models are prone to performance changes (“drift”) over time due to shifts in case-mix or input features.^2,3^ Sudden changes in health care utilization during the SARS-CoV-2 pandemic may have impacted the performance of clinical predictive models.^4^

## METHODS

We evaluated the performance over time of a machine learning, EHR-based mortality prediction algorithm currently used in clinical practice. The algorithm was validated to predict 180-day mortality among outpatients with cancer at medical oncology practices within a large academic cancer center in southeastern Pennsylvania.^5^ Since January 2019, this algorithm has been used as part of an intervention to prompt serious illness conversations among individuals with >10% risk score of 180-day mortality.^6^ Model features are listed at https://github.com/pennsignals/eol-onc and were derived using demographic, comorbidity, and laboratory values within 180 days prior to the encounter. Risk scores are generated on the Thursday prior to each medical oncology encounter and reflect absolute predicted percentage-point mortality risk.

We analyzed mortality risk scores associated with 237,336 in-person and telemedicine medical oncology encounters between January 2019 and December 2020. Descriptive analyses compared encounter-level characteristics in the following periods: January 2019–February 2020 (“pre-pandemic”), March–May 2020 (“early pandemic”, representing the early impact of stay-at-home orders), and June–December 2020 (“later pandemic”, representing longer-term impact).

## PRINCIPAL FINDINGS

Interrupted time series analyses that controlled for pre-pandemic trends evaluated the impact of the pandemic on absolute 180-day mortality risk scores and the monthly percentage of encounters classified as high-risk, using March-May 2020 as a washout period. Relative to the pre-pandemic period, the later pandemic period was characterized by a 6.8-percentage-point decrease (28.2% vs. 35.0%) in high-risk encounters (p<0.001 for level change, **Figure 1A**). There was a corresponding 2.2 absolute percentage-point decrease in predicted mortality risk in the pre-pandemic vs. later-pandemic period. (13.2% vs. 11.0%, p<0.001 for level change, **Figure 1B**).

**Figure 1.**
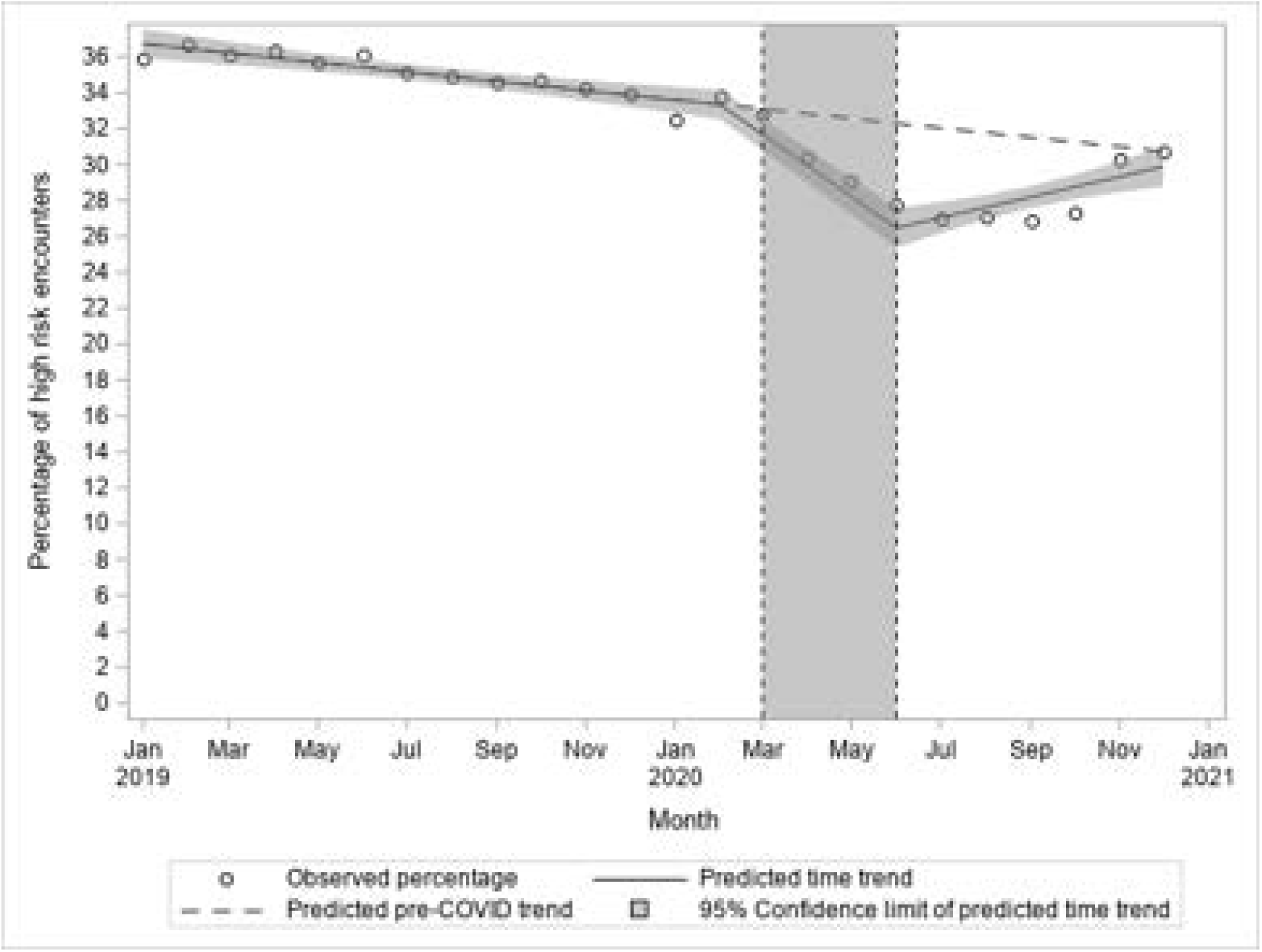
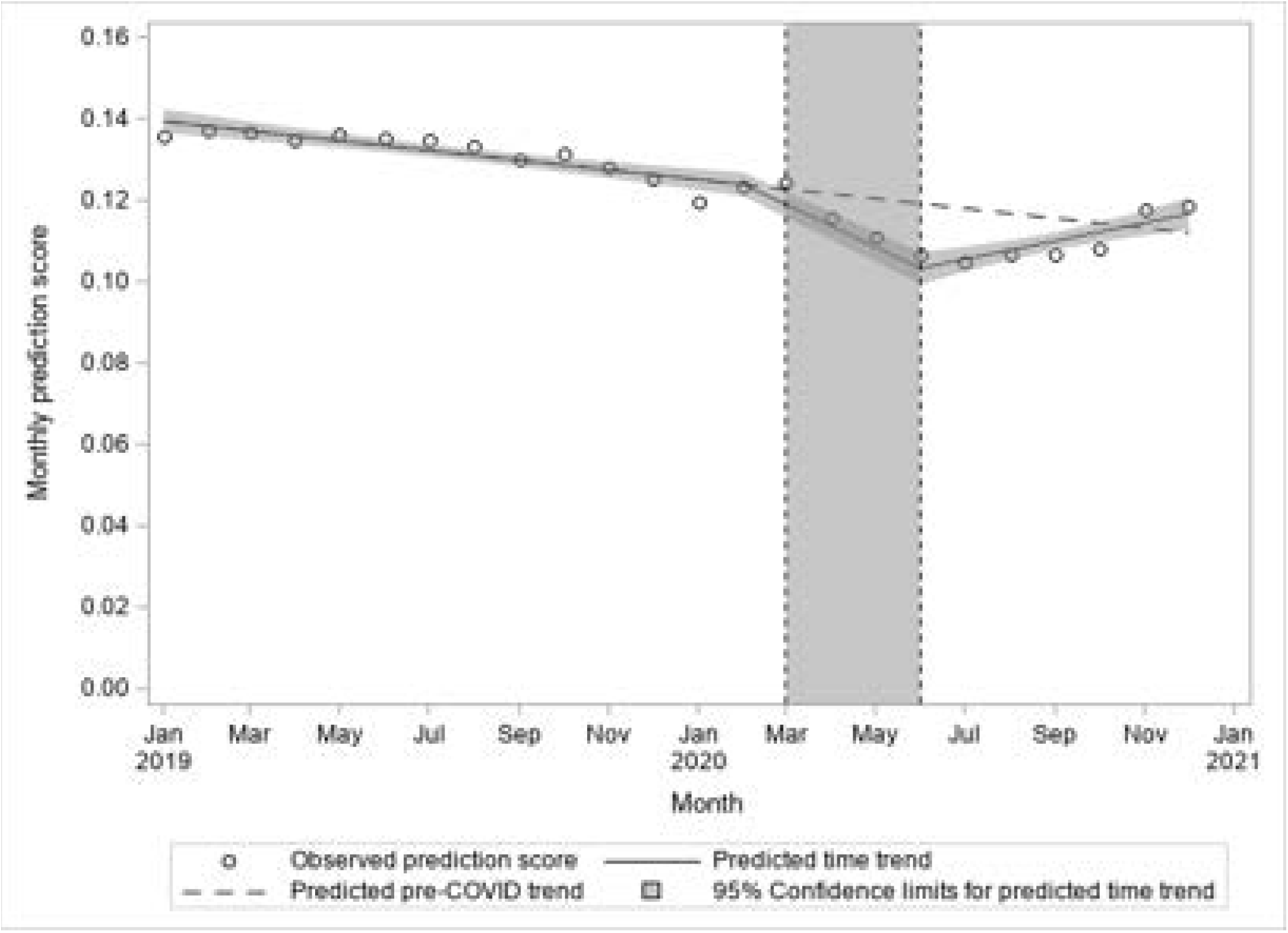
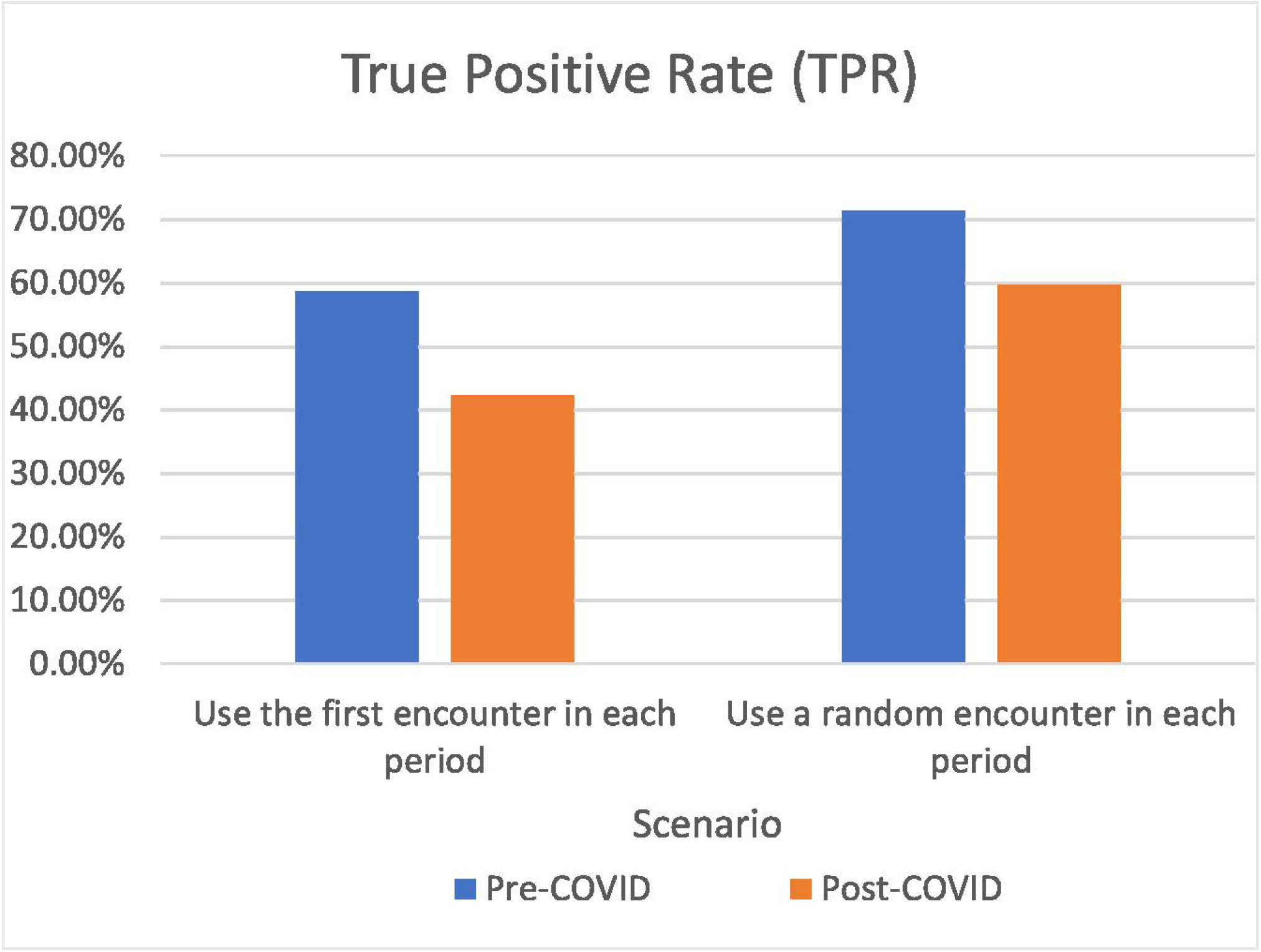
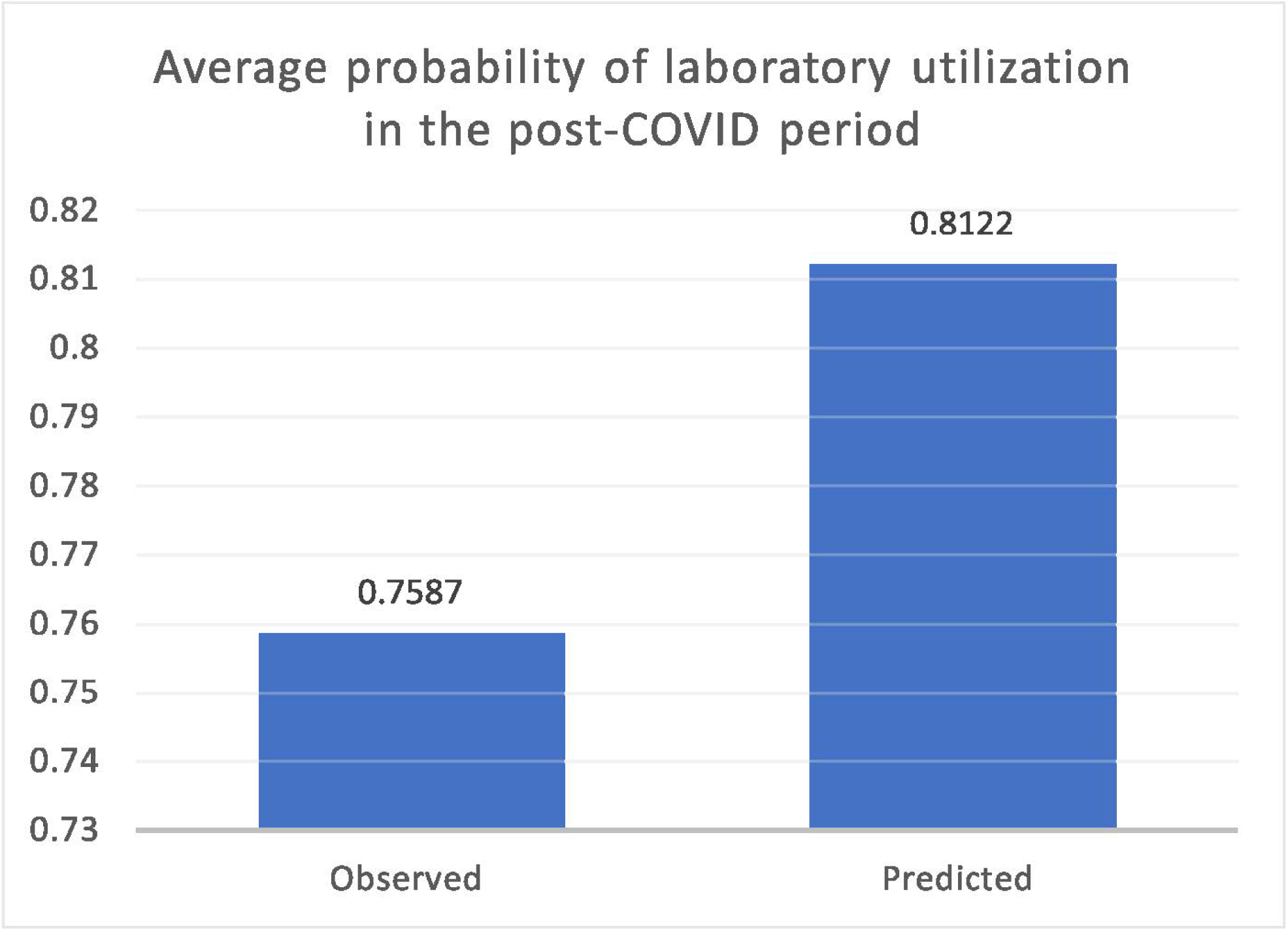
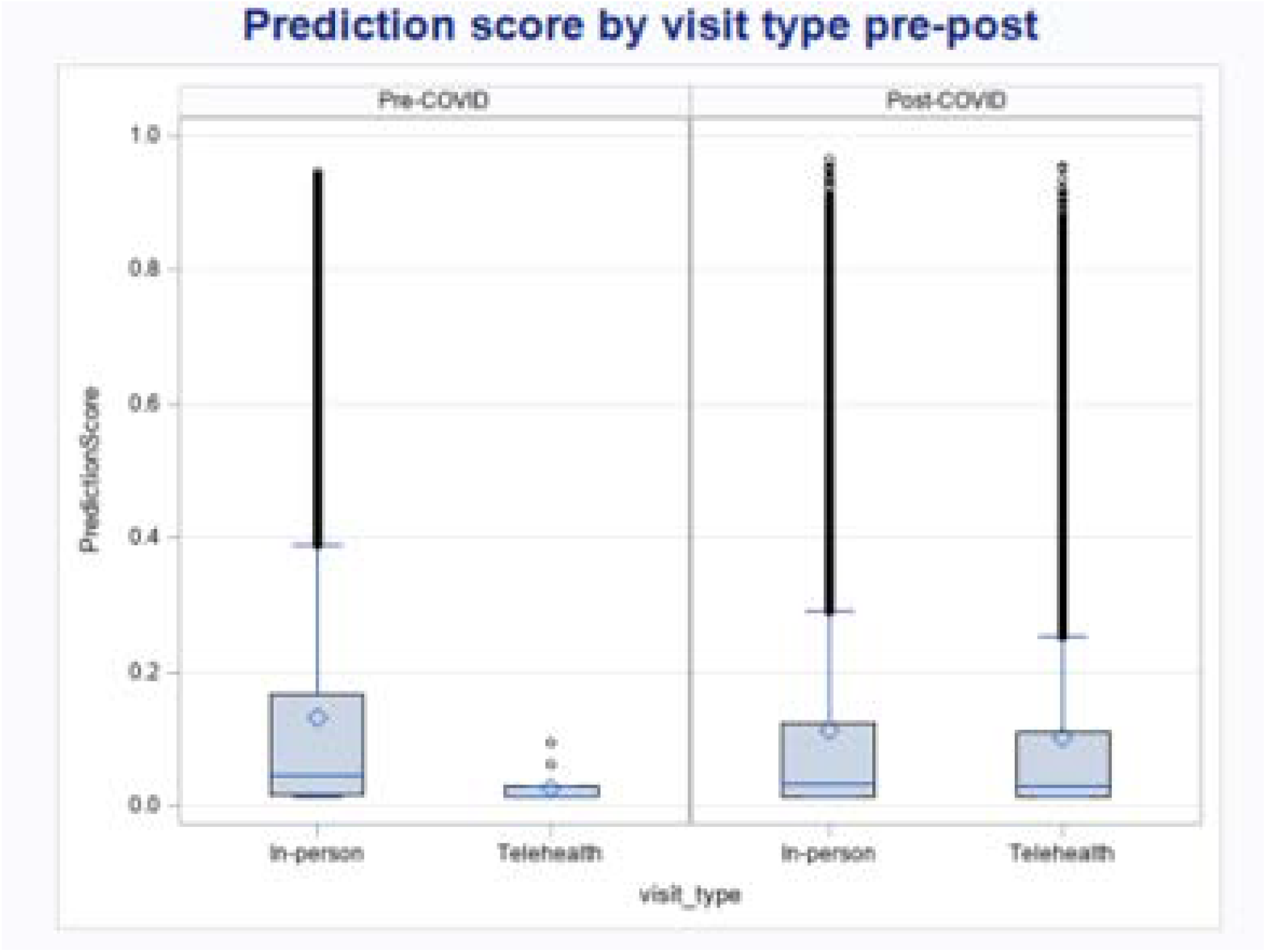
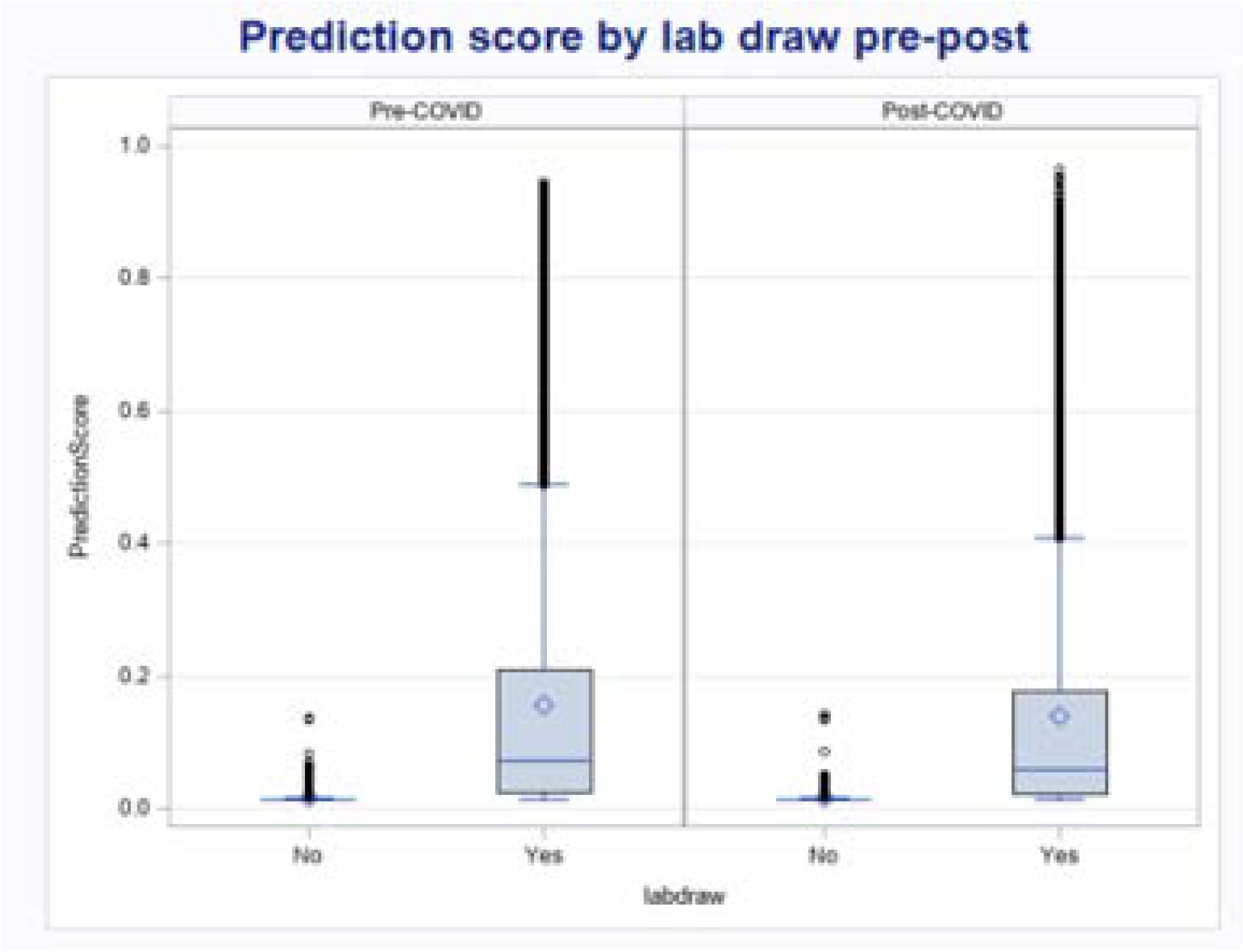
Performance drift in the prediction model during the SARS-CoV-2 pandemic. Interrupted time series analyses of SARS-CoV-2 pandemic impacts on **(A)** percentage of high-risk encounters, and **(B)** absolute predicted 180-day mortality risk. Circles represent monthly observed averages across all medical oncology encounters. Solid lines reflect time trends in the pre-pandemic (January 2019–February 2020), early pandemic (March–May 2020), and later pandemic (June–December 2020) periods, with 95% confidence intervals surrounding each time trend in shaded grey. **(C)** True positive rate of model prediction pre- vs. later pandemic. Both scenarios showed a decrease in the TPR. **(D)** Comparison of observed and predicted laboratory utilization in the later pandemic period. The predicted laboratory utilization was 0.81, while the observed laboratory utilization was 0.76. **(E)** Prediction score by visit type in pre- vs. later pandemic. The mean prediction score in the pre-pandemic period among the in-person visits was 0.13 (IQR: 0.02-0.17), while among telehealth visits, the mean prediction score was 0.03 (IQR: 0.01-0.03). In the later pandemic period, the mean prediction score among in-person visits was 0.11 (IQR: 0.01-0.12), among telehealth visits was 0.12 (0.02-0.14). **(F)** Prediction score by laboratory utilization in pre- vs. later pandemic. The mean prediction score in the pre-pandemic period among the encounters with laboratory utilization was 0.16 (IQR: 0.03-0.21), while among the encounters without any laboratory utilization, the mean prediction score was 0.015 (IQR: 0.013-0.014). In the later pandemic period, the mean prediction score among the encounters with labs was 0.14 (IQR: 0.02-0.18), among the encounters without labs was 0.015 (0.012-0.014).

Because we hypothesized that pandemic-related utilization declines would result in underprediction of mortality risk, true positive rate (TPR) was the primary performance metric. TPR was calculated based on the total number of predicted high-risk encounters and observed all-cause 180-day mortality from the encounter date, based on the 10% threshold used in practice.^6^ Mortality risk scores from each patient’s first encounter in the pre-pandemic and later pandemic periods were used to calculate TPR, using the bootstrap method with 1,000 resamples to generate confidence intervals.^7, 8^ As a sensitivity analysis, we calculated TPR using a random encounter in each period. TPR decreased from 58.8% (95% CI 55.8%, 61.9%) in the pre-pandemic period to 42.3% (95% CI: 36.8%, 47.8%) in the later pandemic period; the sensitivity analysis was consistent (TPR 71.5% [69.1%, 73.9%] vs. 59.8 [54.8%, 64.9%]) (**Figure 1C**).

To identify potential mechanisms of performance drift, we first compared demographic characteristics, which identified differential numbers of lab utilization and telemedicine as primary difference in pre- vs. later pandemic. Age, race, average patient encounters per month, insurance type, comorbidity, laboratory values, and overall mortality were similar across the three time periods (**Table 1**). Compared to the pre-pandemic period, the early and later pandemic periods had higher proportions of telemedicine encounters (20.0% and 26.4% vs 0.01%) and encounters with no preceding laboratory draws (19.8% and 24.1% vs. 17.7%). To further investigate whether laboratory utilization in the later pandemic period was lower than expected, we trained a LASSO regression model based on pre-pandemic data to predict laboratory utilization in the later pandemic period. In the later pandemic period, 81.2% of encounters were predicted to be associated with laboratories, whereas only 75.9% (51,431/67,786) of encounters had associated laboratory utilization that was observed (**Figure 1D**). Finally, to investigate the potential contribution of higher telemedicine encounters and encounters with no preceding laboratory draws to performance drift, we examined mortality risk scores associated with and without telemedicine encounters or laboratory visits. In the later pandemic period, telemedicine encounters (mean predicted risk score 10.3%) and encounters with no preceding laboratory draws (1.5%) were associated with lower predicted risk score than in-person encounters (11.2%, p<0.001) or encounters with preceding laboratory draws (14.0%, p<0.001), respectively (**Figure 1E-F**).

**Table 1:** Baseline characteristics of medical oncology encounters

|  | **Overall** | **Pre-pandemic** | **Early pandemic** | **Late pandemic** |
| --- | --- | --- | --- | --- |
| **Number of encounters, N (%)** | 237336 (100.00) | 141969 (59.82) | 27582 (11.62) | 67785 (28.56) |
| **Number of unique patients, N (%)** | 34666 (100.00) | 27609 (79.64) | 12702 (36.64) | 21961 (63.35) |
| **Monthly encounter rate (Total patient visits per month) (SD)** | 1.47 (0.88) | 1.50 (0.94) | 1.45 (0.82) | 1.42 (0.78) |
| **Prediction Score per encounter, mean (SD)** | 0.12 (0.18) | 0.13 (0.18) | 0.12 (0.17) | 0.11 (0.17) |
| Telemedicine | 0.11 (0.16) | 0.03 (0.03) | 0.12 (0.16) | 0.10 (0.16) |
| In-person | 0.13 (0.18) | 0.13 (0.18) | 0.12 (0.17) | 0.11 (0.17) |
| No preceding labs | 0.02 (0.01) | 0.02 (0.01) | 0.01 (0.01) | 0.02 (0.01) |
| Preceding labs | 0.15 (0.19) | 0.16 (0.19) | 0.14 (0.18) | 0.14 (0.18) |
| **Observed 6-month mortality rate (%)** | 7.0 | 7.4 | 6.3 | 6.4 |
| **Age, mean (SD)** | 61.92 (13.86) | 61.91 (13.74) | 61.95 (13.81) | 61.94 (14.12) |
| **Age group, N (%)** |  |  |  |  |
| <55 years old | 62605 (26.38) | 37305 (26.28) | 7295 (26.45) | 18005 (26.56) |
| 55-64 years old | 64806 (27.31) | 39407 (27.76) | 7419 (26.90) | 17980 (26.53) |
| 65-74 years old | 72553 (30.57) | 43041 (30.32) | 8587 (31.13) | 20925 (30.87) |
| 75 and over | 37372 (15.75) | 22216 (15.65) | 4281 (15.52) | 10875 (16.04) |
| **Marital status, N (%)** |  |  |  |  |
| Married | 155676 (65.59) | 92860 (65.41) | 18306 (66.37) | 44510 (65.66) |
| Not married | 81612 (34.39) | 49087 (34.58) | 9269 (33.61) | 23256 (34.31) |
| Missing | 48 (0.02) | 22 (0.02) | 7 (0.03) | 19 (0.03) |
| **Race, N (%)** |  |  |  |  |
| Non-Hispanic White | 178794 (75.33) | 107305 (75.58) | 20735 (75.18) | 50754 (74.87) |
| Hispanic Latino/White | 2834 (1.19) | 1690 (1.19) | 328 (1.19) | 816 (1.20) |
| Non-Hispanic Black | 36438 (15.35) | 21691 (15.28) | 4179 (15.15) | 10568 (15.59) |
| Hispanic Latino/Black | 1072 (0.45) | 667 (0.47) | 119 (0.43) | 286 (0.42) |
| Other | 13262 (5.59) | 7873 (5.55) | 1632 (5.92) | 3757 (5.54) |
| Unknown | 4936 (2.08) | 2743 (1.93) | 589 (2.14) | 1604 (2.37) |
| **Insurance, N (%)** |  |  |  |  |
| Commercial | 26748 (11.27) | 15767 (11.11) | 3059 (11.09) | 7922 (11.69) |
| Medicaid | 12589 (5.30) | 7530 (5.30) | 1412 (5.12) | 3647 (5.38) |
| Medicare | 193417 (81.50) | 115766 (81.54) | 22592 (81.91) | 55059 (81.23) |
| Self-Pay | 943 (0.40) | 515 (0.36) | 142 (0.51) | 286 (0.42) |
| Missing | 3639 (1.53) | 2391 (1.68) | 377 (1.37) | 871 (1.28) |
| **Visit type, N (%)** |  |  |  |  |
| In-person | 213943 (90.14) | 141958 (99.99) | 22069 (80.01) | 49916 (73.64) |
| Telehealth | 23393 (9.86) | 11 (0.01) | 5513 (19.99) | 17869 (26.36) |
| **Elixhauser comorbidity count, mean (SD)** | 2.34 (1.89) | 2.36 (1.88) | 2.33 (1.87) | 2.30 (1.93) |
| **Encounters with no preceding laboratory values, N (%)** | 46942 (19.8) | 25119 (17.7) | 5469 (19.8) | 16354 (24.1) |
| **Lab value, mean (SD)** |  |  |  |  |
| Albumin | 4.34 (22.00) | 4.33 (23.07) | 4.31 (20.10) | 4.38 (20.36) |
| Hemoglobin | 12.10 (1.75) | 12.09 (1.76) | 12.13 (1.72) | 12.10 (1.73) |
| Calcium | 9.17 (0.49) | 9.18 (0.51) | 9.16 (0.46) | 9.17 (0.45) |
| White blood cell | 7.21 (9.08) | 7.16 (8.15) | 7.13 (8.35) | 7.34 (11.00) |
| Total bilirubin | 0.60 (0.62) | 0.61 (0.62) | 0.58 (0.52) | 0.61 (0.68) |
| Alkaline phosphatase | 91.27 (77.04) | 92.47 (81.25) | 90.85 (70.47) | 88.93 (70.20) |

## CONCLUSIONS

During the SARS-CoV-2 pandemic period, the performance of a machine learning prognostic algorithm used to prompt serious illness conversations in clinical practice declined substantially. The algorithm underidentified patients at high predicted risk of mortality in the initial months of the pandemic period. Towards the end of 2020, prediction scores began to normalize, presumably due to resumption of routine health utilization. Declines in encounters associated with laboratory draws and increases in telemedicine utilization – both potentially spurred by pandemic-related stay-at-home orders and patient fears of exposure in health settings – may have disproportionately contributed to lower performance. Results were directionally consistent in a sensitivity analysis.

Performance drift of predictive algorithms has been conceptually described in certain settings where characteristics of training and validation populations differ.^2,3^ This is one of the first studies to show algorithm performance drift due to SARS-CoV-2 pandemic-related utilization shifts; this drift extended well into 2020. Two potential mechanisms of performance drift are dataset shift or natural algorithm performance drift. We show that scores dropped sharply after the initial SARS-CoV-2 period, which would be atypical for algorithm performance drift. Furthermore, demographic, clinical severity, and cancer-specific characteristics remained largely similar across time periods. Rather, a shift in the underlying utilization patterns of the cohort during the pandemic, resulting in a database shift, likely contributed to algorithm inaccuracy. A similar study published in 2021 found that predictive model alerts increased by 43% during the SARS-CoV-2 pandemic, even though this model was not trained in the SARS-CoV-2 era.^9^ Cancellation of elective surgeries and higher than average patient acuity in the underlying patients contributed to this. Our study adds to this and similar literature by suggesting that changes in the frequency of alerts caused by pandemic-related utilization shifts may be associated with decreased accuracy overall. Predictive or risk-adjustment algorithms that use input values from EHR or claims data should be interpreted with caution, as pandemic-related decreases in utilization may impact performance. Health systems, payers, and clinicians should consider retraining EHR- or claims-based predictive algorithms in the post-pandemic era.

Limitations of this study include its single-institution setting and inability to account for unmeasured contributors to changes in underlying risk. In particular, it is possible that the distribution of other unmeasured markers of patient acuity, including performance status, cancer burden, or other factors, may have contributed to fewer alerts during the pandemic period. While we relied on Social Security and institutional data to measure the outcome of death, and while overall rates of death were similar across periods, it is possible that we did not capture all deaths during the later pandemic period. However, because fewer high-risk alerts were generated in the later pandemic period, it is likely that an increase in the outcome of death would have resulted in a further decrease in the sensitivity of the algorithm.

## POLICY IMPLICATIONS

While some studies have short evidence of changes in scores from predictive models^9^, ours is the first to examine changes in algorithm performance, measured by the decrease in the TPR during the pandemic. Our study strongly argues for considering retraining of models with training data during the lengthy SARS-COV-2 pandemic period, given likely utilization shifts during the pandemic. This is also likely to affect other performance models, including risk stratification and risk-adjustment algorithms used by large payers. Alternatively, other methods such as reinforcement learning can enhance clinical applicability by accounting for changes in the underlying dataset.^10^

## DETAILED METHODS

### Data sources

The study cohort was extracted from Clarity, a database that contains structured data elements of individual EHR data for patients treated at the University of Pennsylvania Health System (UPHS). The EHRs contained patient demographic characteristics, comorbidities, laboratory results, and utilization data. Mortality data was derived from internal administrative data, the EHR, and the Social Security Administration Death Master File, matched to UPHS patients by social security number and date of birth.

### Ethics

The University of Pennsylvania Institutional Review Board approved this study with a waiver of informed consent, classifying this study as quality improvement.

### Participants

Patients were eligible if they were 18 years or older and had an encounter at one of 18 medical or gynecologic oncology clinics within the UPHS between January 2^nd^, 2019, and Dec 31^st^, 2020. All encounters were associated with a mortality risk prediction generated prior to the appointment. Eligible practices included a large tertiary practice, in which clinicians subspecialize in one cancer type (e.g., lymphoma), and 17 general oncology practices, in which clinicians usually treat multiple cancer types. Benign hematology, genetics, and survivorship visits were excluded. 360,727 encounters were eligible after these exclusions.. Encounters with the same contact serial number (CSN) were combined as one distinct encounter including all identified comorbidity conditions. After the combination, 237,336 encounters were included in the analytical sample.

### Predictive Algorithm

The mortality risks of patients were derived from a gradient boosting machine learning algorithm (GBM) designed to predict 180-day mortality among outpatients with cancer. 559 structured EHR features collected at UPHS were used to train this algorithm. The 180-day prospective predictions were generated once a week on Thursdays. Even during the pandemic period, most appointments were scheduled prior to the previous Thursday, and thus scheduling additions or subtractions after the prediction was made were rare. Detailed descriptions of the ML algorithm were described in previous publications.^5,8^ The overall AUC of this algorithm was 0.89 (95% CI, 0.88-0.90), and disease-specific AUC ranged from 0.74 to 0.96 in a prospective validation.^5^

### Outcomes

The primary outcome of the algorithm was 180-day mortality from the time of the encounter. Mortality data was derived from internal administrative data, the EHR, and the Social Security Administration Death Master File.

### Features

Variables used in the algorithm have been previously published and are provided in **Supplementary Table 1**.

## Statistical analysis

### Interrupted Time Series Analysis

An interrupted time series (ITS) model was used to evaluate the changes in the absolute 180-day mortality risk scores and the mortality percentage of encounters classified as high-risk before and after the pandemic, using March-May 2020 as a washout period. The purpose of introducing the washout period in the analysis was to allow the pandemic-related stay-at-home orders to take effect in patient behaviors as well as clinical practices. The model structure is as followed,

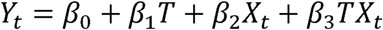

where *T* represents the time elapsed since the start of the study period with the unit representing the frequency with which observations were taken (i.e., month in current study), *X*_*t*_ is a dummy variable indicating the pre-pandemic period (coded in 0) or the later pandemic period (coded in 1), *Y*_*t*_ is the outcome at time *t*. Monthly average prediction score or monthly percentage of high-risk encounters was used in current study. represents the baseline prediction score at *T* =0, *β*_1_ is interpreted as the change in the outcome associated with a time unit increase (representing the underlying pre-pandemic trend), *β*_2_ is the level change following the pre-pandemic period, and *β*_3_ indicates the slope change in the later pandemic period (using the interaction between time and pandemic period: *TX*_*t*_.

The level change in the predicted mortality from pre-pandemic to later pandemic was a decrease of 0.02 (p<.0001), the slope of prediction score was −0.001 (p<.0001) in the pre-pandemic period and 0.003 (p<.0001) in the later pandemic period. The level change in the percentage of high-risk encounters from pre-pandemic to later pandemic was a decrease of 6.7 percent, the slope of percentage of high-risk encounters was −0.26 (p<.0001) in the pre-pandemic period and 0.83 (p<.0001) in the later pandemic period.

### True Positive Rate Comparison

Patients who appeared only in either the pre-pandemic or later pandemic periods were selected. 1,000 bootstrap samples were generated based on this population. Two scenarios were considered in the analysis to calculate the TPR, 1) using the first encounter per patient in each period, or 2) using a random encounter per patient in each period. Encounters flagged with “High-risk” was counted to be compared with the total observed number of patients who died in 6 months from the appointment date. The TPR was calculated in pre-pandemic and later pandemic separately to illustrate whether there was a drift in the model performance.

### Mechanisms of Performance Drift

To compare observed vs. predicted laboratory utilization, a LASSO model was used to predict the laboratory utilization in the later pandemic period. Pre-pandemic period data was used to train the model, predictors included in the model were baseline demographic characteristics (i.e., age, race, marital status, insurance coverage) and comorbidity conditions. Then the modeled output was applied to the later pandemic period to predict the laboratory utilization, including all visit types. The analysis was performed in RStudio.

## Data Availability

All data produced in the present work are contained in the manuscript

## Notes

## Acknowledgements

Thank you to Jay Fein for assistance with manuscript preparation.

## Competing Interests

Dr. Parikh reports receiving grants from Humana, the National Institutes of Health, Prostate Cancer Foundation, National Palliative Care Research Center, Conquer Cancer Foundation, and Veterans Administration; personal fees and equity from GNS Healthcare, Inc. and Onc.AI; personal fees from Cancer Study Group, Thyme Care, Humana, and Nanology; honorarium from Flatiron, Inc. and Medscape; and serving on the board (unpaid) of the Coalition to Transform Advanced Care, all outside the submitted work.

## Funding Information

This work was supported by a grant (to RBP) from the National Cancer Institute (K08-CA-26341).

## Data Access

Drs. Parikh and Zhang had access to the data and vouch for the accuracy of the results.

## Notes

### Competing Interest Statement

The authors have declared no competing interest.

### Funding Statement

This study was funded by NIH/National Cancer Institute K08CA263541-01 grant

### Author Declarations

IRB of the University of Pennsylvania gave ethical approval for this work

